# Suicide Attempt Risk in Autism: A National EHR Study of 2.3 Million Individuals

**DOI:** 10.64898/2026.07.15.26358168

**Authors:** Monika Baker, Wai-yin Lam, Mihai Virtosu, Nina de Lacy

## Abstract

**Background:** Suicide attempts (SA) are elevated in individuals with autism spectrum disorder (ASD), but population-level data characterizing how SA prevalence varies across demographic and clinical subgroups — at the scale and granularity needed to inform evidence-based risk stratification — have been largely unavailable. This study examines SA prevalence across sex, age group, psychiatric comorbidity type, and substance use disorder subtype in the largest real-world ASD cohort to date.

**Methods:** We conducted a retrospective cross-sectional analysis using Epic Cosmos electronic health record data from 2,311,171 individuals with ASD identified by International Classification of Diseases, Tenth Revision, Clinical Modification (ICD-10-CM) codes, spanning 2016–2025. SA prevalence was calculated with Wilson score 95% confidence intervals. Modified Poisson regression with robust variance estimation was used to estimate adjusted prevalence ratios (aPRs) for sex, age group, and psychiatric comorbidity. Unadjusted prevalence ratios (uPRs) were calculated separately for individual comorbidity types and substance use disorder subtypes.

**Results:** Overall SA prevalence was 1.7% (38,160 individuals). Females showed higher SA prevalence than males (2.9% vs. 1.2%; aPR 1.61, 95% CI 1.35–1.92). SA prevalence peaked in the 15–24 age group overall (aPR 5.14, 95% CI 3.62–7.29), with sex-stratified analyses revealing that females peaked earlier (15–24 years; aPR 4.33, 95% CI 4.23–4.43) than males (25–34 years; aPR 6.08, 95% CI 5.05–7.31) — a sex-specific divergence in the timing of peak SA prevalence not previously documented in ASD. Having at least one psychiatric comorbidity was associated with a 30-fold higher SA prevalence (aPR 30.56, 95% CI 26.03–35.89), with the effect stronger in females (aPR 36.54, 95% CI 31.04–43.01) than males (aPR 27.77, 95% CI 22.65–34.06). Among comorbidity subtypes, substance-related disorders showed the highest crude SA prevalence (16.9%; uPR 134.20, 95% CI 67.68–266.10). Subtype-level characterization revealed SA prevalence ranging from 19.7% to 24.3% across all five substance use disorder subtypes examined, with stimulant use disorder showing the highest unadjusted prevalence ratio of any subtype (uPR 196.11, 95% CI 100.63–382.20).

**Conclusion:** SA prevalence in ASD is markedly elevated relative to the general population and varies meaningfully by sex, age, and comorbidity profile in clinically important ways. Females carry a disproportionate SA burden relative to males, with peak vulnerability arriving earlier in adolescence; males peak later in young adulthood and remain at elevated risk into midlife. Psychiatric comorbidity — particularly substance use disorders — is associated with the largest relative elevations in SA prevalence. These population-level estimates are directly applicable to EHR-based risk stratification models and can inform the development of ASD-specific clinical decision support tools that concentrate surveillance and intervention on those at demonstrably elevated risk, rather than applying uniform approaches across a heterogeneous population.

## 1 Introduction

Autism Spectrum Disorder (ASD) is a continuum of complex neurodevelopmental disorders influenced by various environmental and genetic factors, with an etiology that remains incompletely understood (1). While ASD has significant phenotypic heterogeneity (2), it is characterized by deficits in socialization and restricted and repetitive behaviors (3). ASD is associated with multiple comorbidities including attention deficit hyperactivity disorder (ADHD), mood disorders, Obsessive-Compulsive Disorder (OCD), and schizophrenia (4,5). Beyond these comorbid conditions, persons with ASD face elevated rates of suicide attempts (SA) relative to the general population (6). A 2024 meta-analysis estimated that individuals with ASD face a 2.85-fold higher risk of death by suicide compared to the general population (7). Pooled prevalence estimates from recent meta-analyses indicate that between 15–24% of individuals with ASD report a lifetime history of SA (6,8,9). The elevated SA burden in ASD is thought to reflect a confluence of ASD-specific vulnerabilities, including chronic social isolation, difficulties communicating distress, and the psychological costs of masking autistic traits (10,11).

Beyond overall prevalence, the demographic and clinical correlates of SA in ASD — including variation by sex, age, and psychiatric comorbidity — remain incompletely characterized (9,6), in part because much of the existing literature conflates suicide attempts with suicidal ideation, self-harm, or suicidal behaviors broadly (6,9,12,13,14) — outcomes that are conceptually and clinically distinct, with evidence that their associated factors differ meaningfully (15,16). Studies that aggregate these outcomes risk obscuring the specific correlates of SA and limiting the clinical utility of findings for identifying individuals at greatest risk (17). This is especially evident in the literature on sex differences in SA risk, where findings vary considerably depending on which outcome is examined (18,19). Males and females with ASD show similar risk of death by suicide (18), while more recent large population-based studies have identified higher SA risk in females (8,18,20), a pattern that has only emerged as research in this area has grown (19). Autism has historically been characterized as a predominantly male condition, with diagnosis rates and much of the foundational research skewed toward males (18,21). As diagnostic recognition of ASD in females has improved over the past two decades, with female diagnosis rates increasing steadily and females still consistently diagnosed later than males (21), prior estimates of sex differences in SA risk are likely underestimated in older studies conducted when female ASD diagnoses were less common (19,21).

Age is an important demographic correlate of SA risk in ASD, with adolescence and young adulthood consistently identified as a period of heightened vulnerability reflecting the confluence of increased social demands, diagnostic transitions, and reduced structured support during this developmental period (22,23). However, the ASD suicide literature has been heavily skewed toward youth, with research on middle-aged and older adults with ASD accounting for only 0.4% of indexed autism research — a significant gap given emerging evidence that suicidality remains elevated and may even increase with age in individuals with ASD (24,25,26). Population-level, age-stratified SA prevalence data across the full lifespan therefore remain limited, and whether age patterns of risk differ by sex in ASD has not been characterized at scale (9,6).

Psychiatric comorbidity is among the most consistently identified correlates of SA in ASD, with any co-occurring psychiatric disorder associated with elevated SA risk across multiple population-based studies and systematic reviews (9,12,20). Mood disorders represent the most extensively studied comorbidity in this context — depressive symptoms are among the most frequently and consistently identified correlates of suicidality in ASD, though existing evidence largely pertains to suicidal ideation rather than suicide attempts specifically, reflecting the broader measurement heterogeneity in this literature (9,22,27). Co-occurring psychotic disorders have similarly been associated with markedly elevated SA risk in ASD, a relationship thought to reflect compounding vulnerability from both conditions given the well-established link between psychosis and suicidality in the general population (20,28). Trauma and stressor-related disorders warrant particular attention in the ASD context given that individuals with ASD are disproportionately exposed to traumatic experiences — including bullying, victimization, and adverse childhood events — and PTSD has been identified as one of the most strongly associated comorbidities with SA in ASD specifically (11,20). Eating disorders similarly confer markedly elevated SA risk in the general population — with individuals with anorexia nervosa 18 times more likely to die by suicide relative to age- and sex-matched peers — yet their specific association with SA in ASD has received limited empirical attention despite meaningful phenotypic overlap between ASD and eating disorders (12,29). Substance use disorders have been identified as among the strongest comorbidity-associated correlates of SA in ASD (20), yet existing studies have largely examined substance use disorders as a broad category without characterizing risk across specific subtypes such as stimulant, cannabis, opioid, and alcohol use disorders — limiting the granularity of findings available to inform clinical decision-making (12,20).

The burden of SA in ASD has important clinical implications that remain incompletely addressed, as evidence-based prevention strategies and validated assessment approaches specific to ASD are scarce (9,11). Universal suicide risk screening — the application of standardized screening tools to all individuals in a clinical setting regardless of risk — has been examined as one response to this burden, but evidence for its effectiveness remains limited (30,31). The U.S. Preventative Services Task Force (USPSTF) has concluded that evidence is insufficient to recommend for or against universal suicide risk screening in the adult population, citing in part inadequate evidence on the accuracy of available screening tools (30,31). Standard suicide screening tools have historically not been validated for use in ASD populations, where communication differences and atypical symptom presentation complicate standard assessment (32,33). Moreover, clinicians report significantly lower self-efficacy in assessing suicide risk in individuals with ASD compared to clients without ASD (33). These limitations suggest that universal screening approaches alone are insufficient to address the elevated SA burden in ASD (11,33,34).

What is needed instead is a risk stratification approach — defined as the process of classifying patients according to their health risks and projected care needs, enabling clinical resources and monitoring to be directed toward those at highest risk rather than applied uniformly across all patients (35). Unlike universal screening, which aims to identify any individual who may be at risk, risk stratification leverages population-level data on demographic and clinical characteristics to identify which subgroups carry the greatest burden, supporting a more targeted and resource-efficient approach to prevention (35,36). In the context of suicide prevention, EHR-based risk stratification approaches have demonstrated particular promise in general psychiatric and veteran populations — machine learning models applied to structured EHR data have markedly outperformed clinician-led risk assessments, and programs such as the VA Recovery Engagement and Coordination for Health-Veterans Enhanced Treatment (REACH-VET) initiative have demonstrated that targeting enhanced outreach to the highest-risk tier of patients can meaningfully reduce documented suicide attempts (36,37,38). However, existing EHR-based suicide risk models have been developed primarily in general psychiatric and veteran populations, and the clinical features that predict SA risk in ASD differ meaningfully from those in the general population. ASD-specific factors including trauma symptoms, sensory sensitivities, and distinctive comorbidity patterns are important predictors of SA risk in individuals with ASD that would not be captured by general population models (11,39). Developing effective risk stratification approaches for ASD therefore requires population-level prevalence data characterizing how SA risk is distributed across demographic and clinical subgroups specific to this population — data that have been largely unavailable at the scale and granularity needed (6,9). While previous studies have explored the relationship between psychiatric diagnosis and SA in ASD, much of this work has been limited by small sample sizes or single-country datasets (8,9,12,20), and the role of specific substance use disorder subtypes in conferring differential SA risk has not been characterized at population scale.

The present study addresses this need using a large national electronic health record dataset comprising over 2.3 million individuals with ASD — far exceeding the sample sizes of prior population-based studies examining SA in this population, including a Danish population-based cohort of 6,584 individuals with ASD (20), a Swedish national register study of 54,168 individuals with ASD (8), and a Canadian population-based matched cohort of 75,926 individuals with ASD (13). We characterize SA prevalence when stratified by sex, age group, and psychiatric comorbidity type, with particular attention to substance use disorder subtypes — demographic and clinical dimensions that have not been simultaneously characterized in ASD and that are directly relevant to identifying high-risk subgroups for targeted clinical intervention (11,39). Through this characterization, we provide a population-level descriptive foundation needed to inform evidence-based suicide risk stratification in ASD — establishing which subgroups carry the greatest SA burden and supporting more targeted, efficient, and clinically meaningful approaches to suicide prevention in this high-risk population.

## 2 Materials and Methods

### 2.1 Study Design and Data Source

We conducted a retrospective, cross-sectional study using deidentified national EHR data from the Epic Cosmos data warehouse (40). Epic Cosmos aggregates EHRs from more than 2,133 hospitals, 48,800 clinics, and 304 million unique patients, representing geographically and demographically diverse populations across inpatient, outpatient, and emergency settings. Participating health systems agree to contribute their data, which is pooled and de-identified at source. Each patient has a unique identifier, and their inpatient and outpatient charts are integrated into a single comprehensive record that moves with them among constituent healthcare systems. Cosmos data are curated through a multi-stage process managed jointly by Epic and participating health systems. Each site securely transmits its data after mapping local values to standardized vocabularies before transfer. Epic conducts a comprehensive validation to verify data parity with the source systems, corrects mapping errors, and assesses completeness before approving data for inclusion. All workflows were conducted within the Cosmos Data Science Virtual Machine (DSVM) environment, which prohibits the export of any line-level or patient-level data to ensure compliance with data privacy and security standards. The analytic period spanned January 1, 2016, through December 31, 2025.

### 2.2 Study Population

The study population included patients aged five years or older with a lifetime diagnosis of autism spectrum disorder (ASD). ASD diagnoses were identified using the following ICD-10-CM codes: F84.0 (Autistic disorder), F84.5 (Asperger’s syndrome), F84.8 (Other pervasive developmental disorders), and F84.9 (Pervasive developmental disorder, unspecified). Individuals were stratified by sex (female, male) and age group (5–14, 15–24, 25–34, 35–44, 45–64, and 65+ years). Persons whose sex was neither male nor female were excluded. Psychiatric comorbidities were classified using ICD-10-CM codes and included eating disorders, mood disorders, schizophrenia and psychotic disorders, substance-related disorders, and trauma and stressor-related disorders (Supplementary Table 1). Substance-related disorders were further stratified by subtype: alcohol use disorder, opioid use disorder, cannabis use disorder, stimulant use disorder, and other substances (Supplementary Table 1). Individuals with multiple psychiatric diagnoses were included in each relevant comorbidity category.

### 2.3 Outcome Definition

The primary outcome was a recorded suicide attempt during the study period, identified using ICD-10-CM codes for intentional self-harm (Supplementary Table 2). Administrative coding of suicide attempts using ICD-10-CM codes is known to undercount true prevalence, as sensitivity for identifying nonfatal suicidal behaviors in administrative data is low and varies substantially across settings and populations (41). SA prevalence was calculated as the number of unique patients with at least one recorded suicide attempt divided by the total number of patients in each stratum, expressed as a percentage with 95% confidence intervals calculated using the Wilson score method (42).

### 2.4 Statistical Analysis

Descriptive statistics were used to characterize the study population and calculate crude prevalence of suicide attempt across all strata. Modified Poisson regression with robust variance estimation was used to calculate adjusted prevalence ratios (aPRs) and 95% confidence intervals, with history of suicide attempt as the binary outcome. This approach was selected over logistic regression as prevalence ratios are the more appropriate measure of association in cross-sectional studies with binary outcomes, providing more direct and interpretable estimates than odds ratios (43,44). Three models were fit: (1) an overall model including sex, age group, and presence of at least one psychiatric comorbidity as covariates; (2) sex-stratified models to assess whether associations differed between females and males; and (3) separate unadjusted models for each psychiatric comorbidity type and substance use disorder subtype compared to no comorbidity. Comorbidity-specific prevalence ratios are presented unadjusted as the aggregated nature of the data precluded simultaneous adjustment for multiple psychiatric comorbidities, given that individuals may carry more than one diagnosis. Reference categories were male sex, age group 5–14, and no psychiatric comorbidity. Statistical significance was defined as p < 0.05. All analyses were conducted using Python (version 3.13) with the statsmodels library (45). Cell counts of 10 or fewer were suppressed in accordance with Epic Cosmos data privacy policies and excluded from analyses.

### 2.5 Ethics

This study was determined to not meet the definitions of Human Subjects Research by the University of Utah Institutional Review Board (IRB_00202177) and was therefore exempt from informed consent requirements.

### 2.6 Data Availability Statement

The data used in this study are derived from the Epic Cosmos data warehouse and are not publicly available due to data privacy and security restrictions inherent to the Cosmos Data Science Virtual Machine (DSVM) environment. Researchers interested in accessing Epic Cosmos data should contact Epic Systems or their participating institution. The analytic code used in this study is available from the de Lacy lab’s GitHub (delacylab/ASD_Suicide_Prevalence_EHR_Data).

## 3 Results

### 3.1 Sample Characteristics

A total of 2,311,171 individuals with ASD were identified in the Epic Cosmos database during the study period (2016–2025), of whom 629,895 (27.3%) were female and 1,681,276 (72.7%) were male. Overall, 38,160 individuals (1.7%) had at least one recorded suicide attempt during the study period. The age distribution of the sample was as follows: 5–14 years (44.1%), 15–24 years (29.5%), 25–34 years (15.8%), 35–44 years (5.6%), 45–64 years (3.6%), and 65+ years (1.4%). A total of 646,643 individuals (28.0%) had at least one recorded psychiatric comorbidity, with females more likely to have a comorbidity (36.8%) than males (24.7%). Full sample characteristics are presented in Table 1. SA prevalence by sex and age group is examined in the sections that follow.

**TABLE 1.** Sample characteristics of individuals with ASD, stratified by sex. Demographic and clinical characteristics of 2,311,171 individuals with ASD identified in the Epic Cosmos database (2016–2025). Counts and percentages are reported for females, males, and the total sample.

| Characteristic | Female N (%) | Male N (%) | Total N (%) |
| --- | --- | --- | --- |
| Total patients | 629,895 (27.3%) | 1,681,276 (72.7%) | 2,311,171 (100%) |
| Suicide attempt | 18,158 (2.9%) | 20,002 (1.2%) | 38,160 (1.7%) |
| Age group |  |  |  |
| 5–14 | 257,403 (40.9%) | 762,828 (45.4%) | 1,020,231 (44.1%) |
| 15–24 | 180,519 (28.7%) | 500,320 (29.8%) | 680,839 (29.5%) |
| 25–34 | 106,511 (16.9%) | 259,078 (15.4%) | 365,589 (15.8%) |
| 35–44 | 45,171 (7.2%) | 84,883 (5.0%) | 130,054 (5.6%) |
| 45–64 | 29,531 (4.7%) | 52,650 (3.1%) | 82,181 (3.6%) |
| 65+ | 10,760 (1.7%) | 21,517 (1.3%) | 32,277 (1.4%) |
| Comorbidity |  |  |  |
| At least one | 232,074 (36.8%) | 414,569 (24.7%) | 646,643 (28.0%) |
| None | 397,821 (63.2%) | 1,266,707 (75.3%) | 1,664,528 (72.0%) |

### 3.2 Suicide Attempt Prevalence by Sex

Females with ASD had considerably higher SA prevalence than males (2.9% vs 1.2%; 18,158 vs 20,002 recorded suicide attempts), and this difference remained significant after adjustment for age group and psychiatric comorbidity (aPR 1.61, 95% CI 1.35–1.92, p = 1.07e-07), representing a 61% higher prevalence in females; overall adjusted prevalence ratios are presented in Figure 1. Sex-stratified analyses examining age and comorbidity effects separately are presented in Table 2 and described in the sections below.

**FIGURE 1.**
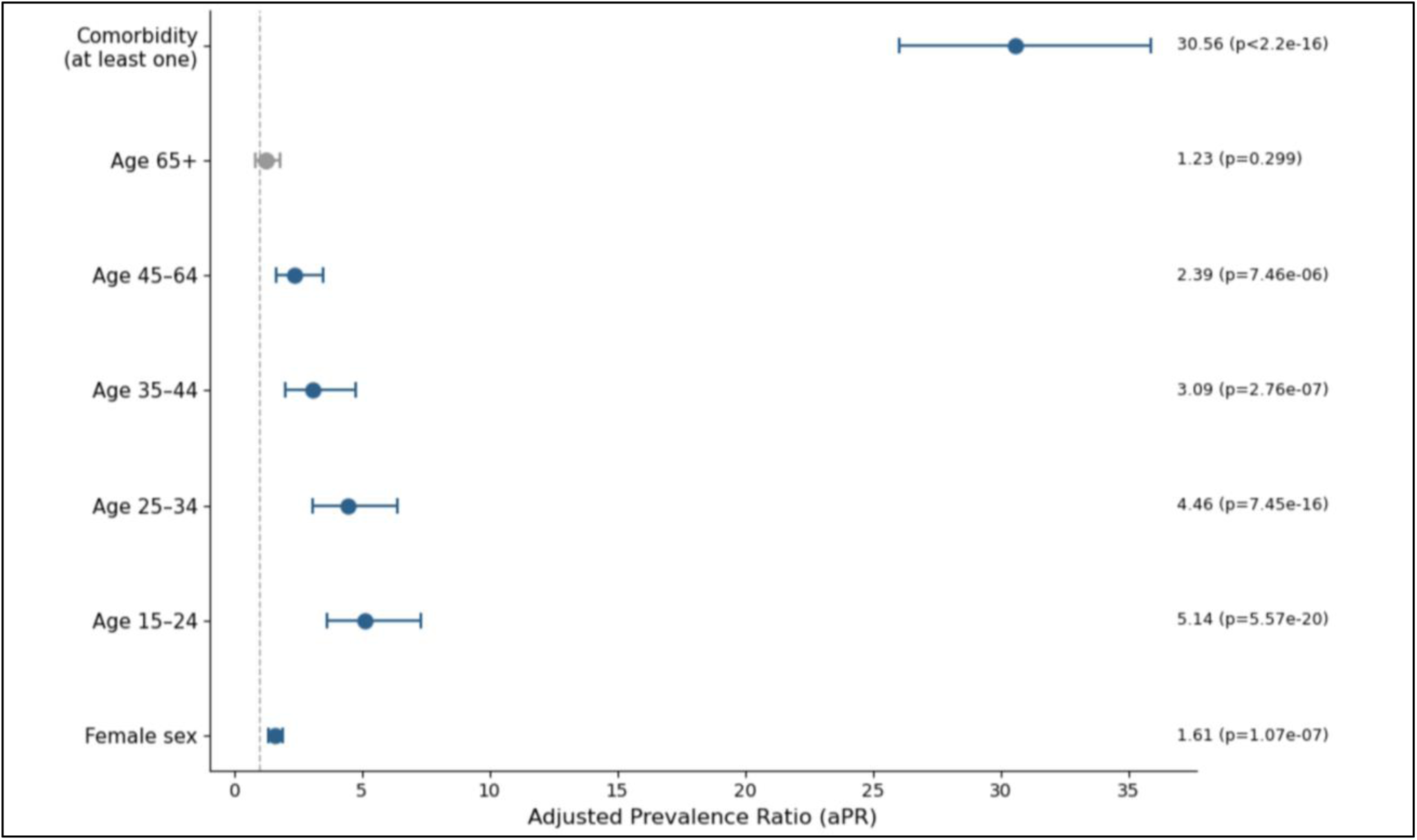
Adjusted prevalence ratios (aPRs) for suicide attempt in individuals with autism spectrum disorder: overall model. Points represent aPRs with 95% confidence intervals from modified Poisson regression with robust variance estimation. Reference categories: male sex, age group 5–14 years, no psychiatric comorbidity. Gray point indicates non-significant association (p = 0.299).

**TABLE 2.** Adjusted prevalence ratios (aPRs) for suicide attempt in individuals with ASD: sex-stratified models. Sex-stratified adjusted prevalence ratios from modified Poisson regression with robust variance estimation, with age group 5–14 years and no psychiatric comorbidity as reference categories.

| Variable | Female<br>aPR | Female<br>95% CI | Female<br>p-value | Male<br>aPR | Male<br>95% CI | Male<br>p-value |
| --- | --- | --- | --- | --- | --- | --- |
| **Age group** |  |  |  |  |  |  |
| 5–14 (ref) | — | — | — | — | — | — |
| 15–24 | 4.33 | 4.23–4.43 | < 2.20e-16 | 5.74 | 4.81–6.85 | < 2.20e-16 |
| 25–34 | 3.02 | 2.92–3.12 | < 2.20e-16 | 6.08 | 5.05–7.31 | < 2.20e-16 |
| 35–44 | 1.92 | 1.88–1.95 | < 2.20e-16 | 4.60 | 3.84–5.51 | < 2.20e-16 |
| 45–64 | 1.58 | 1.55–1.62 | < 2.20e-16 | 3.35 | 2.81–3.99 | < 2.20e-16 |
| 65+ | 0.80 | 0.78–0.82 | < 2.20e-16 | 1.71 | 1.44–2.04 | 1.52e-09 |
| **Comorbidity** |  |  |  |  |  |  |
| At least one | 36.54 | 31.04–43.01 | < 2.20e-16 | 27.77 | 22.65–34.06 | < 2.20e-16 |
| None (ref) | — | — | — | — | — | — |

*ASD = autism spectrum disorder. aPR = adjusted prevalence ratio. CI = confidence interval. ref = reference category. Modified Poisson regression with robust variance estimation (HC3). Cell counts of ≤10 suppressed per Epic Cosmos data privacy policies and excluded from analysis*

### 3.3 Suicide Attempt Prevalence by Age

SA prevalence varied markedly across age groups, peaking in the 15–24 age group and declining progressively with advancing age. Crude SA prevalence was 0.23% in the 5–14 reference group, rising to 2.79% in the 15–24 age group, 3.12% in the 25–34 age group, 2.60% in the 35–44 age group, 2.09% in the 45–64 age group, and 0.99% in the 65+ age group. After adjustment, all age groups showed significantly elevated SA prevalence relative to the 5–14 reference group except the 65+ age group: 15–24 (aPR 5.14, 95% CI 3.62–7.29, p = 5.57e-20), 25–34 (aPR 4.46, 95% CI 3.10– 6.42, p = 7.45e-16), 35–44 (aPR 3.09, 95% CI 2.01–4.76, p = 2.76e-07), 45–64 (aPR 2.39, 95% CI 1.63–3.50, p = 7.46e-06), and 65+ (aPR 1.23, 95% CI 0.83–1.80, p = 0.299). Full adjusted prevalence ratios by age group are presented in Table 3.

**TABLE 3.** Adjusted prevalence ratios (aPRs) for suicide attempt in individuals with ASD: overall model. Adjusted prevalence ratios from modified Poisson regression with robust variance estimation, including sex, age group, and presence of at least one psychiatric comorbidity as covariates. Reference categories: male sex, age group 5–14 years, no psychiatric comorbidity.

| Variable | aPR | 95% CI | p-value |
| --- | --- | --- | --- |
| **Sex** |  |  |  |
| Female | 1.61 | 1.35–1.92 | 1.07e-07 |
| Male (ref) | — | — | — |
| **Age group** |  |  |  |
| 5–14 (ref) | — | — | — |
| 15–24 | 5.14 | 3.62–7.29 | 5.57e-20 |
| 25–34 | 4.46 | 3.10–6.42 | 7.45e-16 |
| 35–44 | 3.09 | 2.01–4.76 | 2.76e-07 |
| 45–64 | 2.39 | 1.63–3.50 | 7.46e-06 |
| 65+ | 1.23 | 0.83–1.80 | 2.99e-01 |
| **Comorbidity** |  |  |  |
| At least one | 30.56 | 26.03–35.89 | < 2.20e-16 |
| None (ref) | — | — | — |

### 3.4 Suicide Attempt Prevalence by Age and Sex

Age patterns differed meaningfully by sex. Among females, crude SA prevalence peaked in the 15–24 age group (5.40%) and declined progressively across older groups (25–34: 4.60%; 35–44: 3.19%; 45–64: 2.62%; 65+: 1.23%), with an adjusted peak at 15–24 years (aPR 4.33, 95% CI 4.23–4.43). Among males, crude SA prevalence was lower across all age groups, peaking at 25–34 years (2.51%) rather than 15–24 years (1.84%), with an adjusted peak similarly at 25–34 years (aPR 6.08, 95% CI 5.05–7.31). Notably, females aged 65+ showed a significantly lower SA prevalence compared to the 5–14 reference group (aPR 0.80, 95% CI 0.78–0.82), while males aged 65+ remained at elevated risk (aPR 1.71, 95% CI 1.44–2.04, p = 1.52e-09). Sex-stratified age results are presented in Table 2 and Figure 2.

**FIGURE 2.**
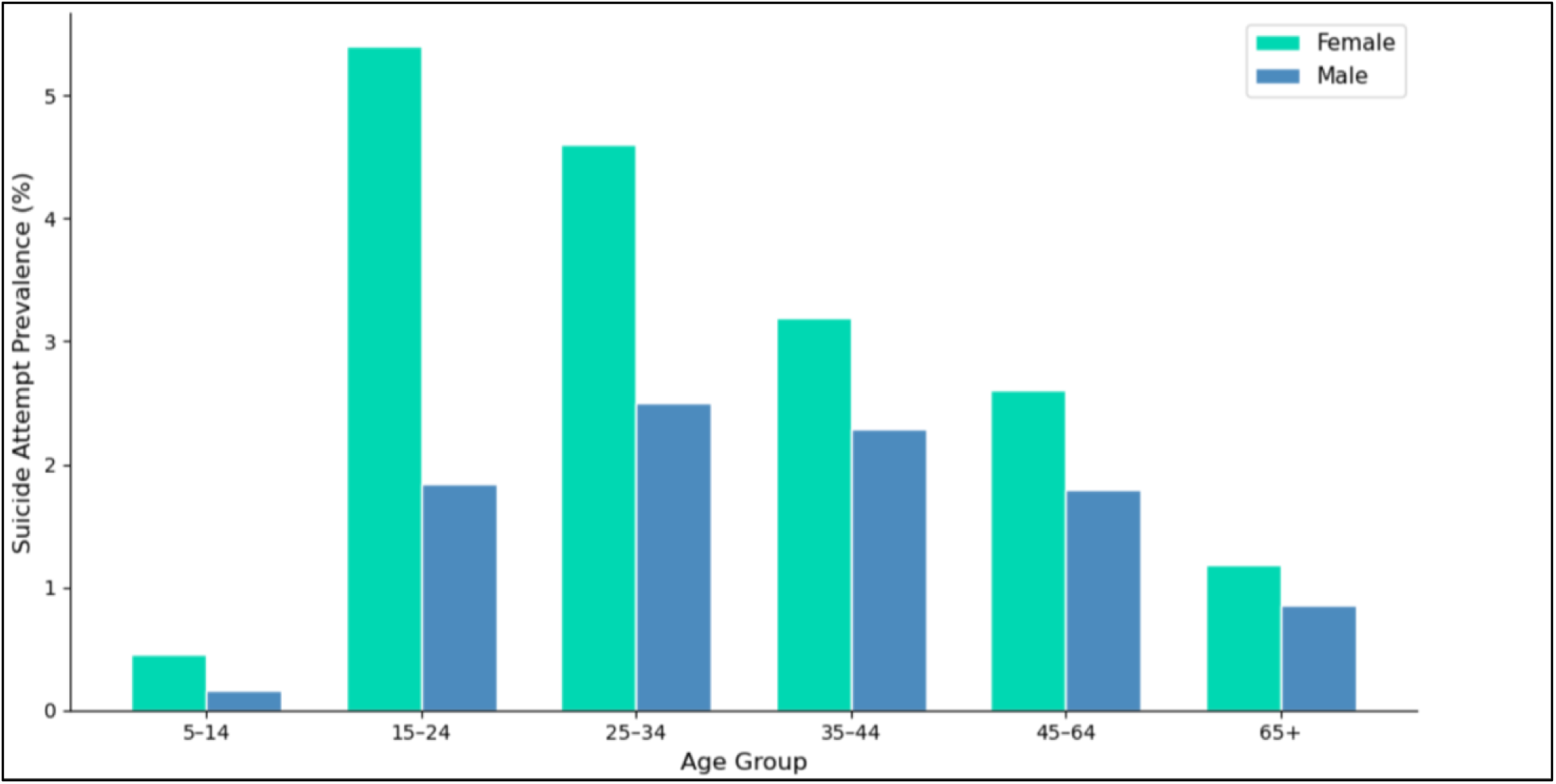
Suicide attempt prevalence by sex and age group in individuals with autism spectrum disorder. Bars represent crude prevalence (%) of recorded suicide attempt within each age group, stratified by sex.

### 3.5 Suicide Attempt Prevalence by Psychiatric Comorbidity

Having at least one psychiatric comorbidity was associated with markedly higher SA prevalence compared to having no comorbidity. Among individuals with at least one psychiatric comorbidity, crude SA prevalence was 7.49% compared to 0.21% in those with no comorbidity, representing a 30-fold difference after adjustment (aPR 30.56, 95% CI 26.03–35.89, p < 2.20e-16). Full adjusted prevalence ratios are presented in Table 3.

SA prevalence varied substantially across comorbidity types (Table 4). Substance-related disorders were associated with the highest crude SA prevalence (16.9%) and the highest unadjusted prevalence ratio among comorbidity categories (uPR 134.20, 95% CI 67.68–266.10, p < 2.20e-16), followed by schizophrenia and psychotic disorders (crude prevalence 11.8%; uPR 93.89, 95% CI 47.21–186.72, p < 2.20e-16). Trauma and stressor-related disorders (8.2%), mood disorders (7.4%), and eating disorders (5.0%) showed lower but still considerably elevated crude prevalences; unadjusted prevalence ratios for all comorbidity types are presented in Table 5. Substance-related disorders were further stratified by subtype; subtype-specific prevalence estimates and unadjusted prevalence ratios are presented in the following section.

**TABLE 4.**
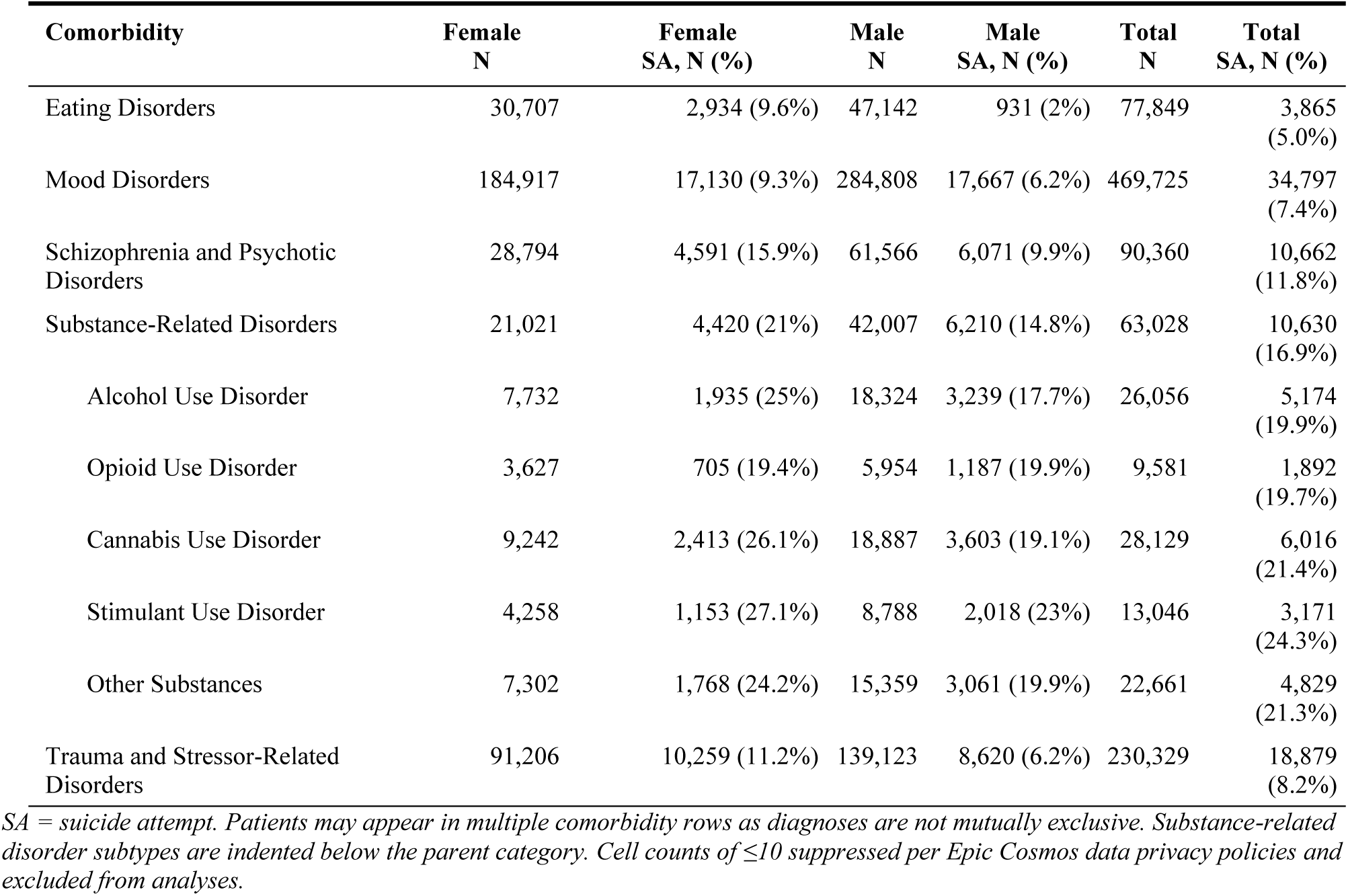
Prevalence of suicide attempt by psychiatric comorbidity, stratified by sex. Sample sizes and crude suicide attempt prevalence for each psychiatric comorbidity category and substance use disorder subtype, reported for females, males, and the total sample. Substance use disorder subtypes are indented below the parent category. Individuals with multiple diagnoses are included in each relevant category.

**TABLE 5.** Unadjusted prevalence ratios (uPRs) for suicide attempt by psychiatric comorbidity type and substance use disorder subtype in individuals with ASD. Each comorbidity type and substance use disorder subtype was compared separately against individuals with no psychiatric comorbidity as the reference group. uPRs are presented unadjusted, as the aggregated nature of the data precluded simultaneous adjustment for multiple comorbidities.

| Comorbidity | uPR | 95% CI | p-value |
| --- | --- | --- | --- |
| **Psychiatric comorbidity type** |  |  |  |
| Eating Disorders | 39.78 | 12.12–130.60 | 1.25e-09 |
| Mood Disorders | 58.95 | 29.35–118.40 | < 2.20e-16 |
| Schizophrenia and Psychotic Disorders | 93.89 | 47.21–186.72 | < 2.20e-16 |
| Substance-Related Disorders | 134.20 | 67.68–266.10 | < 2.20e-16 |
| Trauma and Stressor-Related Disorders | 65.22 | 29.32–145.09 | < 2.20e-16 |
| **Substance use disorder subtype** |  |  |  |
| Alcohol Use Disorder | 158.71 | 80.18–314.16 | < 2.20e-16 |
| Opioid Use Disorder | 174.59 | 88.56–344.19 | < 2.20e-16 |
| Cannabis Use Disorder | 170.70 | 87.30–333.78 | < 2.20e-16 |
| Stimulant Use Disorder | 196.11 | 100.63–382.20 | < 2.20e-16 |
| Other Substances | 177.89 | 91.29–346.64 | < 2.20e-16 |

The effect of psychiatric comorbidity differed by sex. Among females with at least one psychiatric comorbidity, crude SA prevalence was 9.3% compared to 0.45% in females with no comorbidity (aPR 36.54, 95% CI 31.04–43.01, p < 2.20e-16). Among males, crude SA prevalence was 6.2% with comorbidity compared to 0.16% without (aPR 27.77, 95% CI 22.65–34.06, p < 2.20e-16). Sex-stratified comorbidity results are presented in Table 2.

Crude SA prevalence among individuals with at least one psychiatric comorbidity was elevated across all age groups, peaking in the 15–24 age group (7.86% with comorbidity vs 0.26% without) and declining progressively with advancing age. Age-stratified comorbidity prevalence data are presented in Supplementary Table 3.

### 3.6 Suicide Attempt Prevalence by Substance Use Disorder Subtype

Among individuals with substance-related disorders, SA prevalence varied across subtypes. Stimulant use disorder was associated with the highest crude SA prevalence (24.3%) and the highest unadjusted prevalence ratio of any subtype examined (uPR 196.11, 95% CI 100.63–382.20, p < 2.20e-16), followed by other substances (crude prevalence 21.3%; uPR 177.89, 95% CI 91.29– 346.64, p < 2.20e-16), opioid use disorder (19.7%; uPR 174.59, 95% CI 88.56–344.19, p < 2.20e-16), cannabis use disorder (21.4%; uPR 170.70, 95% CI 87.30–333.78, p < 2.20e-16), and alcohol use disorder (19.9%; uPR 158.71, 95% CI 80.18–314.16, p < 2.20e-16). All substance use disorder subtypes showed substantially higher unadjusted prevalence ratios than the broader substance-related disorders category (uPR 134.20). Full subtype-specific results are presented in Table 5.

Crude SA prevalence across all substance use disorder subtypes was highest in the 15–24 age group and declined progressively with advancing age. Stimulant use disorder showed the highest crude SA prevalence in the 15–24 age group (35.6%), followed by opioid use disorder (31.0%), alcohol use disorder (30.1%), other substances (29.1%), and cannabis use disorder (26.7%). Age-stratified subtype prevalence data are presented in Supplementary Table 3.

SA prevalence across subtypes was consistently higher in females than males. Among females, stimulant use disorder showed the highest crude prevalence (27.1%; uPR 178.26, 95% CI 71.39–445.16, p < 2.20e-16), followed by cannabis use disorder (26.1%; uPR 167.56, 95% CI 67.73– 414.54, p < 2.20e-16) and alcohol use disorder (25.0%; uPR 161.36, 95% CI 64.45–403.97, p < 2.20e-16). Among males, stimulant use disorder again showed the highest unadjusted prevalence ratio (uPR 198.75, 95% CI 86.60–456.14, p < 2.20e-16), followed by opioid use disorder (uPR 192.49, 95% CI 82.43–449.47, p < 2.20e-16) and other substances (uPR 183.75, 95% CI 80.23– 420.85, p < 2.20e-16). Sex-stratified subtype results are presented in Supplementary Table 4.

## 4 Discussion

This study characterized suicide attempt prevalence in the largest real-world cohort of individuals with ASD to date, comprising 2,311,171 individuals drawn from the Epic Cosmos database — far exceeding the sample sizes of prior population-based studies, including a Swedish national register study of 54,168 individuals with ASD (8) and a Canadian population-based matched cohort of 75,926 individuals with ASD (13), the latter of which examined self-harm events broadly rather than suicide attempts specifically. This sample size enables stable prevalence estimates across multiple demographic and clinical subgroups simultaneously, stratified by sex, age group, and psychiatric comorbidity — providing the population-level descriptive foundation needed to inform evidence-based suicide risk stratification in ASD. Our findings demonstrate that SA prevalence is elevated relative to the general population in ASD, particularly among those with psychiatric comorbidities, and that sex and age patterns of SA prevalence differ meaningfully between females and males.

The overall SA prevalence of 1.7% in our sample is lower than estimates from previous studies. A recent systematic review reported lifetime SA prevalence of 15.3% and 12-month SA prevalence of 14.1% in ASD (9), while past-year SA prevalence in the general US adult population is approximately 0.8% (46). The lower prevalence observed in the current study likely reflects key methodological differences — our cross-sectional analysis captures SA recorded within a fixed EHR observation window using ICD-10-CM diagnostic codes, rather than self-reported lifetime or 12-month history, which tends to yield higher estimates (6,9). Additionally, administrative coding of SA is known to undercount true prevalence, as ICD-10-CM codes demonstrate limited sensitivity for identifying nonfatal suicidal behaviors and only attempts resulting in a documented clinical encounter within participating health systems are captured (41). Despite these methodological differences, our observed prevalence of 1.7% overall exceeds general population estimates and is consistent with the elevated SA burden documented in prior ASD research (8,9). However, aggregate prevalence figures alone obscure the variation in SA burden across demographic and clinical subgroups. Prevalence rising to 16.9% in those with substance-related disorders and 11.8% in those with schizophrenia and psychotic disorders illustrates the concentration of burden within specific subgroups — and why population-level subgroup characterization is essential for developing targeted risk stratification approaches in ASD (9,47).

### 4.1 Sex Differences in Suicide Attempt Prevalence

Females with ASD have significantly higher SA prevalence than males (2.9% vs. 1.2%; aPR 1.61, 95% CI 1.35–1.92), a finding consistent with an evolving literature in which sex differences in ASD suicidality have only recently come into focus (6,9,18,19). Earlier population-based studies found no significant sex difference in SA prevalence in ASD (48,49), while more recent large population-based studies — including a Danish population-based cohort of 6,584 individuals with ASD (20) and a Swedish national register study of 54,168 individuals with ASD (8) — identified higher SA prevalence in females with ASD than males, a pattern subsequently confirmed in systematic reviews and meta-analyses (6,9,19). The emergence of this sex difference in more recent and larger studies likely reflects improved diagnostic recognition of ASD in females over the past two decades — ASD has historically been characterized as a predominantly male condition, with foundational research skewed toward male presentations and females consistently underdiagnosed or diagnosed later (18,21,50,51), and prior estimates drawn from cohorts with lower female representation are likely to have underestimated sex differences in SA prevalence (18,19). The sex difference in SA prevalence in ASD mirrors the pattern observed in the general population, where females also report higher rates of SA than males (46,52), though the mechanisms associated with elevated SA prevalence in females with ASD are likely distinct from those in the general population, reflecting ASD-specific vulnerabilities — including camouflaging, delayed diagnosis, and elevated psychiatric comorbidity burden — documented across qualitative and quantitative research over the past decade (10,11,19,39,53). It is important to note that this sex difference in SA prevalence does not extend to suicide death — studies of suicide mortality in ASD have consistently found comparable rates between males and females with ASD (8,18) — a divergence that underscores the importance of examining SA and suicide death as conceptually and clinically distinct outcomes, and one that is obscured when suicidal behaviors are aggregated, as has been common in prior research (6,9,14,15,18).

Several converging ASD-specific vulnerabilities may account for the elevated SA prevalence observed in females with ASD relative to males. Camouflaging — the conscious or unconscious suppression of autistic traits to conform to social expectations — is more prevalent in females with ASD than males and is directly associated with suicidality, likely through its links to psychological defeat and internal entrapment (10,19,53,54). Females with ASD face greater gendered social expectations to socialize and conform, generating higher camouflaging demands and the sustained psychological costs of inauthenticity across social contexts (10,19,53). Diagnostic delay compounds this burden — females with ASD are consistently diagnosed later than males on average (21,51,54), resulting in prolonged periods of unrecognized struggle, unmet support needs, and potential misattribution of distress before ASD identification occurs (22,53,54). Elevated psychiatric comorbidity burden in females with ASD further compounds vulnerability: in our sample, 36.8% of females had at least one psychiatric comorbidity compared to 24.7% of males, and the association between comorbidity and SA prevalence was stronger in females (aPR 36.54) than in males (aPR 27.77), consistent with evidence that females with ASD have higher rates of internalizing disorders than males with ASD (9,12,20). These converging vulnerabilities — greater camouflaging demands, later diagnosis, and higher psychiatric comorbidity burden — likely interact to produce the elevated SA prevalence observed in females with ASD relative to males, though the relative contributions of each remain incompletely characterized (11,18).

The age pattern of SA prevalence differs meaningfully between sexes. Among females with ASD, SA prevalence peaks in the 15–24 age group (aPR 4.33, 95% CI 4.23–4.43), while among males it peaks later at 25–34 years (aPR 6.08, 95% CI 5.05–7.31). This sex-specific divergence in the age of peak SA prevalence has not previously been documented at population scale in ASD, as prior studies have generally lacked the sample sizes needed to examine age-by-sex interactions in SA specifically (9,6,18). The earlier peak in females is consistent with the intensification of social demands during adolescence and early adulthood — the period when camouflaging becomes most cognitively and emotionally costly, when undiagnosed females with ASD are most likely to be struggling without adequate support, and when the gap between social expectations and autistic experience may be most acutely felt (19,22,23,53,54). The later peak in males at 25–34 years may reflect the accumulation of stressors associated with post-educational transitions — including high rates of unemployment, difficulties with independent living, and social isolation — that disproportionately affect males with ASD as they enter adulthood (22,27,55). Within the interpersonal theory of suicide, these experiences map onto thwarted belongingness and perceived burdensomeness, both of which have been specifically implicated in ASD suicidality and may accumulate gradually across the transition to adulthood in males rather than manifesting acutely in adolescence (27,55). These sex-specific age patterns suggest that clinical attention in suicide prevention efforts should be directed toward females with ASD in the 15–24 age window and males with ASD across a broader range extending into the mid-twenties and beyond.

The lower SA prevalence in females with ASD aged 65+ relative to the 5–14 reference group (aPR 0.80, 95% CI 0.78–0.82) — in contrast to males aged 65+ who remain at elevated risk (aPR 1.71, 95% CI 1.44–2.04) — is an interesting finding that likely reflects multiple non-mutually-exclusive processes. Selective survival is one plausible contributor: females with ASD who reach older age may represent a subgroup with lower baseline vulnerability, a phenomenon identified as an important consideration in studies of older adults with neurodevelopmental disorders more broadly (56). Cohort effects are likely also relevant — older females with ASD in our sample were far less likely to have received formal ASD recognition during their lifetime given historically low diagnostic rates in females (21,50,51), and this systematic underidentification would suppress documented SA rates through reduced clinical contact and diagnostic coding (25,26,41). The persistently elevated SA prevalence in older males with ASD (aPR 1.71) is consistent with the well-documented vulnerability of older males to suicide-related outcomes in the general population, where social isolation following relationship loss or bereavement, declining physical health, and reduced help-seeking behavior converge to sustain risk into late life (46,52). In ASD, these general population patterns may be compounded by the cumulative burden of lifelong social exclusion and the relative absence of established social networks that might otherwise serve as protective factors in older adulthood — consistent with emerging evidence that suicidality in adults with ASD does not follow the same age-related decline observed in the general population and may even increase with age in some groups (24,25,26).

### 4.2 Age Differences in Suicide Attempt Prevalence

SA prevalence peaks in the 15–24 age group (aPR 5.14, 95% CI 3.62–7.29) and declines progressively with advancing age, though it remains significantly elevated relative to the 5–14 reference group across all age groups except 65+. This overall age pattern is broadly consistent with prior population-based data showing that adolescence and young adulthood represent a period of heightened SA vulnerability in ASD (20,22,23). The mechanisms underlying this peak are likely multifactorial, reflecting the confluence of increasing social demands, diagnostic transitions, loss of structured educational support, and the onset or exacerbation of psychiatric comorbidities that converge during this developmental window (11,22,23). ADHD — among the most common comorbidities in ASD (5) — may be a particularly important contributor to elevated SA risk during adolescence and young adulthood, given that ADHD-associated impulsivity, emotional dysregulation, and executive function deficits have been independently associated with SA risk in adolescent populations, and untreated ADHD in individuals with ASD has been specifically linked to elevated suicidality in late adolescence and early adulthood (5,22,23). Formal support for individuals with ASD often declines during this period — service reductions frequently begin before high school graduation and continue thereafter, creating a window of heightened vulnerability with diminished institutional scaffolding precisely when the demands of adult life are increasing (27,55,57).

The progressive decline in SA prevalence from the 25–34 age group onward — with aPRs of 4.46 (25–34), 3.09 (35–44), and 2.39 (45–64) — is broadly consistent with the general population trend of declining SA prevalence with advancing age (46,52). However, SA prevalence in our ASD sample exceeds general population estimates across all age groups, and individuals with ASD who die by suicide have a mean age at death of 32.4 years — considerably younger than the non-ASD average of 41.8 years — suggesting that suicide risk in ASD extends well into midlife rather than concentrating exclusively in adolescence (58). The persistently elevated SA prevalence in the 35–44 and 45–64 age groups — despite the progressive decline — likely reflects the accumulation of chronic stressors that disproportionately affect adults with ASD across the midlife period, including sustained high rates of unemployment, social isolation, relationship difficulties, and the cumulative burden of untreated or undertreated psychiatric comorbidities (11,27,55). These findings add population-scale evidence to an important but undercharacterized dimension of ASD suicide research — the persistence of elevated risk across the adult lifespan — a gap that has been highlighted given that research on middle-aged and older adults with ASD accounts for only 0.4% of indexed autism research (24,25,26).

The non-significant association in the 65+ age group overall (aPR 1.23, 95% CI 0.83–1.80, p = 0.299) warrants careful interpretation rather than straightforward conclusion that SA risk attenuates in older adults with ASD. Several non-mutually-exclusive explanations are plausible. Systematic underidentification of ASD in older cohorts is likely a major contributor — estimates suggest that approximately nine in ten adults with ASD aged 50 and over in the UK remain undiagnosed or misdiagnosed (26), and this suppression of ASD ascertainment would similarly reduce the likelihood of SA documentation in clinical records (25,41). Survivor bias may also contribute, whereby older individuals with ASD who remain engaged with the healthcare system represent a subgroup with lower baseline vulnerability than those who have died prematurely or disengaged from care (56). The sex-specific breakdown of this finding — with females aged 65+ showing significantly lower SA prevalence than the reference group (aPR 0.80) while males aged 65+ remain at elevated risk (aPR 1.71) — is discussed in the context of cohort effects, diagnostic recognition, and selective survival in Section 4.1 above, and illustrates why the overall 65+ estimate should not be interpreted without this stratification.

Emerging evidence raises important questions about whether SA risk truly attenuates with age in adults with ASD, or whether the apparent decline reflects underascertainment and diagnostic gaps rather than a genuine reduction in vulnerability (24,25). In a cross-sectional study of over 9,900 adults aged 50 and over, depression, anxiety, PTSD, loneliness, and social isolation each mediated the relationship between autistic traits and suicidal ideation, while male sex, depression, PTSD, and social isolation specifically mediated suicidal self-harm (26). The vulnerabilities that drive elevated SA risk during the transition to adulthood in ASD — social exclusion, unemployment, and limited social network formation — do not necessarily resolve with age and may instead accumulate across the lifespan in ways that sustain rather than attenuate risk (26,27,55). The combination of these ASD-specific vulnerabilities with general population risk factors for suicidality in older adulthood — including bereavement, physical health decline, and reduced help-seeking — may sustain SA risk in older adults with ASD at levels that administrative data systematically underestimate (25,52). Taken together, these findings highlight the need for lifespan approaches to suicide prevention in ASD that do not treat adolescence and young adulthood as the exclusive window of risk and point to the importance of targeted surveillance and outreach for middle-aged and older adults with ASD — a group that remains dramatically underrepresented in suicide research and prevention efforts (24,25,26).

### 4.3 Role of Psychiatric Comorbidity in Suicide Attempt Prevalence

Having at least one psychiatric comorbidity is associated with a 30-fold higher SA prevalence in our sample (aPR 30.56, 95% CI 26.03–35.89), far exceeding the SA burden observed in individuals with ASD and no comorbidity. This effect is consistent with — though not directly comparable to — general population data showing that any psychiatric disorder is associated with elevated suicide risk, where a 2022 systematic review and meta-analysis found an OR of 13.1 for suicide associated with any psychiatric disorder (59). The larger relative effect observed in our ASD sample likely reflects the particular challenges that psychiatric comorbidity poses in this population — atypical symptom presentation in ASD can complicate the recognition, diagnosis, and treatment of co-occurring conditions, potentially allowing psychiatric illness to go unidentified or undertreated for longer periods and amplifying its impact on SA risk (9,11,33). This is consistent with prior population-based evidence that psychiatric comorbidity is among the most consistently identified correlates of SA in ASD across multiple datasets and diagnostic categories (8,12,20). Methodological differences limit direct numeric comparison with general population estimates — our prevalence ratios are calculated within the ASD sample relative to those without comorbidity rather than against a non-ASD referent — and should be interpreted in that context.

SA prevalence varies meaningfully across specific comorbidity types, and the pattern of findings across categories provides clinically important granularity beyond the overall comorbidity effect. Among broad comorbidity categories, substance-related disorders are associated with the highest crude SA prevalence (16.9%; uPR 134.20), and their subtype-level characterization — a novel contribution of the current study — is discussed in detail in the following section. Schizophrenia and psychotic disorders are associated with the second highest crude SA prevalence (11.8%; uPR 93.89). The elevated SA prevalence in this group is consistent with prior evidence from ASD populations and with the well-established link between psychosis and suicidality in the general population, where hopelessness, depression, and command hallucinations have been identified as key mediating mechanisms (20,28). Co-occurring psychosis in ASD may represent a particularly high-risk combination, given that ASD-specific vulnerabilities — including difficulties communicating distress and limited social support — may compound the suicide risk already elevated by psychosis alone (12,28). Trauma and stressor-related disorders are associated with an 8.2% crude SA prevalence (uPR 65.22), consistent with evidence that individuals with ASD are disproportionately exposed to traumatic experiences — including bullying, victimization, and adverse childhood events — and that PTSD has been identified as one of the most strongly associated comorbidities with SA in ASD specifically, reflecting the intersection of trauma exposure with ASD-specific emotion regulation difficulties (11,20). Mood disorders show a 7.4% crude SA prevalence (uPR 58.95) and represent the most extensively studied comorbidity in the ASD suicidality literature — depression has been identified as one of the most consistently associated correlates of suicidal thoughts and behaviors in ASD across multiple systematic reviews and meta-analyses, and depressive symptoms are thought to interact with ASD-specific cognitive features such as rigidity and perseveration to amplify SA risk (9,12,22,27). Eating disorders show the lowest crude SA prevalence among comorbidity categories (5.0%; uPR 39.78), though they confer markedly elevated SA prevalence relative to those with no comorbidity, and a striking sex difference — females with ASD and eating disorders show a crude SA prevalence of 9.6% compared to 2.0% in males. This sex disparity is consistent with the well-documented phenotypic and etiological overlap between ASD and eating disorders, particularly anorexia nervosa, which disproportionately affects females and has been associated with elevated SA risk both in the general population and in the context of ASD (12.29).

The association between psychiatric comorbidity and SA prevalence differs by sex — stronger in females with ASD (aPR 36.54) than in males (aPR 27.77), as described in Section 4.1 above. This sex-specific pattern reinforces the broader finding that females with ASD face a compounding burden — higher comorbidity rates, later diagnosis, and a stronger relative impact of comorbidity on SA prevalence — that collectively elevates their SA burden relative to males across the lifespan (12,20,21). The age-stratified comorbidity data further show that crude SA prevalence in those with at least one psychiatric comorbidity peaks in the 15–24 age group (7.86% vs. 0.26% without comorbidity) and declines progressively with age, consistent with the overall age pattern and reinforcing the concentration of comorbidity-associated SA burden during adolescence and young adulthood in ASD (9,22).

### 4.4 Substance Use Disorder Subtypes and Suicide Attempt Prevalence

Substance-related disorders are associated with the highest SA prevalence among all broad comorbidity categories in our cohort (16.9%; uPR 134.20), consistent with prior evidence that substance use disorders are among the most strongly associated comorbidities with SA in ASD (12,20) and with the well-established association between substance use disorders and elevated suicide risk in the general population (47,59). National Medicaid data have documented increasing prevalence of substance use disorder diagnoses among individuals with ASD over time, with cannabis use disorder emerging as the most prevalent subtype in ASD-only populations — a pattern compounded by co-occurring psychiatric disorders including depression (60,61). The current study extends this prior work by characterizing SA prevalence across specific substance use disorder subtypes in individuals with ASD at population scale — a level of granularity that has not previously been reported in this population and that carries direct relevance for targeted clinical monitoring.

All five substance use disorder subtypes examined are associated with SA prevalence ranging from 19.7% to 24.3%, each far exceeding both the overall substance-related disorder category prevalence of 16.9% and the overall ASD cohort prevalence of 1.7%. This convergence of high SA prevalence across all subtypes is consistent with broader general population evidence that alcohol, cannabis, opioids, and stimulants each independently elevate suicide risk through overlapping but distinct pathways — including substance-induced depression, disinhibition of impulse control, pharmacological effects on mood regulation, and the progressive occupational and social decline that frequently accompanies substance use disorder (47,59). In individuals with ASD, these general mechanisms may be compounded by pre-existing ASD-specific vulnerabilities — including difficulties with emotion regulation, limited social support networks, and challenges communicating distress — that reduce capacity to buffer the psychological impact of substance use disorder and may amplify the transition from ideation to attempt (10,11,20).

Stimulant use disorder is associated with the highest crude SA prevalence of any subtype (24.3%; uPR 196.11) in both females (27.1%) and males (23.0%). In the general population, stimulant use — particularly methamphetamine — is associated with elevated suicidality through direct neurobiological effects on dopaminergic and serotonergic systems that impair mood regulation and impulse control, as well as through stimulant-induced mood disorders including depression and dysphoria during withdrawal (62). In individuals with ASD, pre-existing difficulties with emotional regulation — a characteristic feature of ASD that is further amplified by co-occurring ADHD, the most prevalent ASD comorbidity — may compound stimulant-related dysregulation and lower the threshold for suicidal behavior (5,11,22). Cannabis use disorder is associated with high crude SA prevalence in both females (26.1%; uPR 170.70) and males. The association between cannabis use disorder and suicidality is well-documented in the general population, with cannabis use linked to elevated SA prevalence across adolescents and young adults, and females with cannabis use disorder consistently showing higher rates of suicidal ideation and SA than males — a sex difference that is mirrored in our findings and that has been attributed in part to the greater severity of cannabis-associated internalizing symptoms in females, including depression and anxiety (47,63,64). In individuals with ASD, cannabis use may carry elevated risk given the elevated rates of cannabis use disorder documented in this population specifically and the potential for cannabis-induced exacerbation of social withdrawal and mood dysregulation in individuals already vulnerable to these outcomes (11,60,61).

Opioid use disorder is associated with a crude SA prevalence of 19.7% (uPR 174.59). One proposed mechanism linking opioid use disorder to suicidality involves social pain — chronic social exclusion is associated with dysfunction in the endogenous opioid system, which in turn contributes to depression and suicidality; opioid use disorder may emerge partly in response to this social pain while simultaneously deepening the underlying vulnerability (65). In individuals with ASD, where chronic social exclusion is among the most consistent features of the condition across the lifespan, this mechanism may carry particular salience and warrants further investigation (27,55,65). Alcohol use disorder is associated with the lowest SA prevalence among subtypes in our cohort (19.9%; uPR 158.71), though it remains far above both the overall ASD cohort prevalence and general population estimates. The association between alcohol use disorder and suicidality operates through multiple mechanisms including alcohol-induced depression, cognitive constriction, disinhibition of impulse control during acute intoxication, and acceleration of the transition from suicidal ideation to action — all of which may interact with ASD-specific difficulties in emotion regulation to amplify risk (47,59).

The consistently higher SA prevalence in females than males across all SUD subtypes is a novel finding that has not previously been characterized in ASD at population scale. Stimulant use disorder shows the highest crude SA prevalence in both females (27.1%) and males (23.0%), while cannabis use disorder shows the second highest in females (26.1%). This sex pattern across all SUD subtypes is consistent with the broader finding throughout our cohort that females with ASD carry a disproportionate SA burden relative to males, and likely reflects the compounding of substance use disorder with the ASD-specific vulnerabilities documented in Section 4.1 — including camouflaging demands, later diagnosis, and higher psychiatric comorbidity burden — rather than substance use disorder alone (10,19,20). The age-stratified findings further show that SA prevalence peaks in the 15–24 age group across all SUD subtypes — with stimulant use disorder showing the highest crude SA prevalence in this age group (35.6%), followed by opioid use disorder (31.0%), alcohol use disorder (30.1%), other substances (29.1%), and cannabis use disorder (26.7%). This concentration of SUD-associated SA burden during adolescence and young adulthood is consistent with the overall age pattern described in Section 4.2, and reinforces the importance of substance use disorder screening during this developmental window for individuals with ASD — a period when formal support is declining, psychiatric comorbidities are emerging, and social demands are increasing simultaneously (9,22,57). That SA prevalence across SUD subtypes remains elevated across midlife despite progressive decline is consistent with the lifespan vulnerability pattern described in Section 4.2 and underscores that the intersection of substance use disorder and SA risk in ASD is sustained rather than confined to adolescence (20,24).

### 4.5 Limitations

This study has several methodological limitations that should be considered when interpreting the findings. The use of aggregated count data precluded individual-level analysis and simultaneous adjustment for multiple psychiatric comorbidities — as individuals may carry more than one psychiatric diagnosis, comorbidity-specific and substance use disorder subtype prevalence ratios are presented unadjusted and reflect the association of each comorbidity type with SA relative to those with no comorbidity, rather than independent effects after mutual adjustment. Age-stratified comorbidity and substance use disorder subtype estimates are similarly unadjusted and should be interpreted as descriptive rather than causal. The cross-sectional design and fixed observation window (2016–2025) prevented calculation of person-time at risk for individual patients, precluding the estimation of true incidence rates and limiting the ability to draw conclusions about temporal relationships between comorbidity onset and SA.

SA were identified using ICD-10-CM codes recorded within the Epic Cosmos system, and this approach is known to undercount true SA prevalence in administrative data — a rapid review of ICD-10-based surveillance found that sensitivity for identifying nonfatal suicidal behaviors using administrative codes is low, with studies reporting sensitivity ranging from 12% to 86% and substantial undercounting of true SA events (41). Attempts occurring outside of participating health systems, prior to the study period, or not resulting in a clinical encounter would not be captured. Additionally, ICD-10-CM codes do not distinguish between suicide attempts and non-suicidal self-injury in some coding contexts, which may introduce misclassification (41). These limitations are likely to result in underestimation of true SA prevalence across all subgroups examined and may differentially affect older age groups and populations with lower healthcare engagement — including older adults with ASD, who face disproportionate barriers to diagnosis and clinical contact (25,26).

Data on race and ethnicity and intellectual disability were not included in the current analysis, which limits generalizability and the ability to fully characterize the study population. Intellectual disability in particular has a complex and documented relationship with SA risk in ASD — population-based evidence suggests that ASD without co-occurring intellectual disability is associated with higher SA risk than ASD with co-occurring intellectual disability, while the relationship varies by severity of intellectual disability and is further modulated by psychiatric comorbidity (8,11). The absence of intellectual disability data therefore limits the interpretability of our estimates, as SA prevalence in our cohort may reflect a mixture of individuals with and without intellectual disability whose risk profiles differ meaningfully. Race and ethnicity data are similarly important given documented racial and ethnic disparities in suicide-related outcomes in both the general population and in ASD (52). Both variables are captured within the Epic Cosmos infrastructure at the individual level, and their inclusion represents an important priority for future work. Cell counts of 10 or fewer were suppressed in accordance with Epic Cosmos data privacy policies and excluded from analyses, which may have introduced some bias in estimates for smaller subgroups, particularly in older age categories and for less prevalent substance use disorder subtypes.

Finally, while Epic Cosmos is one of the largest real-world clinical datasets available — aggregating EHR data from over 2,133 hospitals across the United States — it captures only individuals who have accessed healthcare within participating systems, and factors influencing healthcare engagement can result in certain groups being underrepresented relative to the broader population (66).

Individuals with ASD who are undiagnosed, have limited healthcare access, or are seen exclusively outside of Cosmos-affiliated institutions would not be captured, and the cohort may therefore skew toward individuals with greater healthcare utilization and more documented clinical contact. These limitations should inform how findings are generalized to the broader ASD population and highlight the importance of complementing EHR-based analyses with data from community and self-report samples.

## 5 Conclusion

Prior population-based studies have established that individuals with ASD face elevated SA risk relative to the general population, but the scale, demographic breadth, and clinical granularity needed to characterize how that risk is distributed across subgroups — and to support evidence-based risk stratification — have been largely unavailable (6,9,12). The present study addresses this gap using a national EHR cohort of over 2.3 million individuals with ASD, providing simultaneously stratified SA prevalence estimates across sex, age, psychiatric comorbidity type, and substance use disorder subtype that prior studies have not been able to generate at this resolution or scale.

The findings reveal a pattern of variation in SA prevalence across demographic and clinical subgroups that is clinically meaningful and that has important implications for how preventive attention should be directed. Psychiatric comorbidity is associated with a 30-fold higher SA prevalence relative to those without comorbidity (aPR 30.56), with the effect stronger in females (aPR 36.54) than in males (aPR 27.77) — consistent with evidence that females with ASD face a compounding burden through camouflaging, diagnostic delay, and higher comorbidity rates that collectively elevates their SA burden beyond what comorbidity alone would predict (10,19,21). Substance-related disorders carry the highest SA prevalence among comorbidity categories, and the subtype-level characterization reported here — with SA prevalence ranging from 19.7% to 24.3% across all five subtypes and stimulant use disorder showing the highest unadjusted prevalence ratio (uPR 196.11) — provides a granularity of clinical information not previously available in ASD (20,62). SA prevalence peaks in the 15–24 age group overall (aPR 5.14) and differs meaningfully by sex — females peaking earlier at 15–24 years (aPR 4.33) and males later at 25–34 years (aPR 6.08) — a sex-specific divergence in the timing of peak SA prevalence that likely reflects distinct developmental pathways and that has direct implications for how clinical attention should be directed across the lifespan (22,27,54). SA prevalence remains elevated well into midlife across all subgroups, and the non-significant association in the 65+ age group overall likely reflects under ascertainment rather than true attenuation of risk — consistent with the well-documented underdiagnosis of ASD in older adults and the known sensitivity limitations of ICD-10-CM coding for nonfatal suicidal behaviors (24,26,41).

These findings are directly relevant to suicide risk stratification in ASD — the process of using population-level data on demographic and clinical characteristics to identify which subgroups carry the greatest burden and to direct preventive resources toward them, rather than applying uniform approaches across a heterogeneous population (35,36). Standard suicide screening tools have not been validated for use in ASD, universal screening has not demonstrated effectiveness in the adult population broadly, and clinicians report lower self-efficacy in assessing suicide risk in individuals with ASD compared to clients without ASD — underscoring the need for stratified, evidence-based approaches that concentrate surveillance and intervention on those at demonstrably elevated risk (30,33,34). EHR-based machine learning models have demonstrated strong clinical utility for suicide risk stratification in psychiatric populations, and extending this approach to ASD requires precisely the kind of population-level descriptive foundation that the current study provides (36,37,67). The subgroup-level SA prevalence estimates reported here — identifying females with ASD, individuals in the 15–24 and 25–34 age windows, and those with co-occurring psychiatric and substance use disorders as the groups carrying the greatest SA burden — provide the empirical grounding needed to develop and validate ASD-specific risk stratification tools (11,39).

Future work should extend this foundation in several directions. Incorporation of intellectual disability status and race and ethnicity data will be essential to characterizing the full heterogeneity of SA risk within ASD and ensuring that stratification approaches are equitable across subpopulations (8,52). Longitudinal analyses examining trajectories of SA risk over time, the temporal relationship between comorbidity onset and SA, and the impact of ASD-specific clinical interventions on SA outcomes would meaningfully complement the cross-sectional prevalence estimates reported here (9,11,34). Community and self-report samples are also needed to reach individuals who do not access healthcare within EHR-affiliated systems — particularly older adults with ASD, who face disproportionate barriers to diagnosis and clinical engagement and whose SA burden is likely underestimated in administrative data (25,26). Taken together, the present findings establish a population-level empirical foundation for more targeted, evidence-based, and clinically meaningful approaches to suicide prevention in ASD — a population carrying a disproportionate and preventable burden of suicidal behavior that has been insufficiently characterized and insufficiently addressed.

## Conflict of Interest

The authors declare that the research was conducted in the absence of any commercial or financial relationships that could be viewed as conflicts of interest.

## Author Contributions

MB: Conceptualization, Formal analysis, Methodology, Software, Validation, Visualization, Writing - original draft, Writing – review & editing. WL: Data Curation, Investigation, Methodology, Resources, Writing - review & editing. MV: Data curation, Formal Analysis, Software, Writing - review & editing. NdL: Conceptualization, Methodology, Project Administration, Supervision, Validation, Writing – review & editing

## Funding

The author(s) received financial support from the Huntsman Mental Health Foundation.

## Acknowledgements

The authors would like to thank David Danks for his contributions to the content of the methodology and the manuscript. The authors acknowledge the use of Claude (Anthropic, San Franciso, CA) for assistance with language editing and manuscript revision. All content was reviewed, verified, and approved by the authors, who take responsibility for the accuracy of the published work.

## Notes

### Competing Interest Statement

The authors have declared no competing interest.

